# Unraveling the Associations Between Voice Pitch and Major Depressive Disorder: A Multisite Genetic Study

**DOI:** 10.1101/2024.10.12.24315366

**Authors:** Yazheng Di, Elior Rahmani, Joel Mefford, Jinhan Wang, Vijay Ravi, Aditya Gorla, Abeer Alwan, Kenneth S. Kendler, Tingshao Zhu, Jonathan Flint

**Affiliations:** CAS Key Laboratory of Behavioral Science, Institute of Psychology, Beijing 100101, China; Department of Psychology, University of Chinese Academy of Sciences, Beijing 100049, China; Department of Computational Medicine, University of California Los Angeles, Los Angeles, CA, USA; Department of Neurology, University of California Los Angeles, Los Angeles, CA, USA; Department of Electrical and Computer Engineering, University of California Los Angeles, Los Angeles, CA, USA; Bioinformatics Interdepartmental Program, University of California Los Angeles, Los Angeles, CA, USA; Virginia Institute for Psychiatric and Behavioral Genetics, Richmond, VA, USA; Department of Psychiatry, Virginia Commonwealth University School of Medicine, Richmond, VA, USA; Department of Psychiatry and Biobehavioral Sciences, Brain Research Institute, University of California Los Angeles, Los Angeles, CA, USA

## Abstract

Major depressive disorder (MDD) often goes undiagnosed due to the absence of clear biomarkers. We sought to identify voice biomarkers for MDD and separate biomarkers indicative of MDD predisposition from biomarkers reflecting current depressive symptoms. Using a two-stage meta-analytic design to remove confounds, we tested the association between features representing vocal pitch and MDD in a multisite case-control cohort study of Chinese women with recurrent depression. Sixteen features were replicated in an independent cohort, with absolute association coefficients (beta values) from the combined analysis ranging from 0.24 to 1.07, indicating moderate to large effects. The statistical significance of these associations remained robust, with P-values ranging from 7.2 × 10^−6^ to 6.8 × 10^−58^. Eleven features were significantly associated with current depressive symptoms. Using genotype data, we found that this association was driven in part by a genetic correlation with MDD. Significant voice features, reflecting a slower pitch change and a lower pitch, achieved an AUC-ROC of 0.90 (sensitivity of 0.85 and specificity of 0.81) in MDD classification. Our results return vocal features to a more central position in clinical and research work on MDD.

## Introduction

Changes in human pitch and tone of speech have been noted as an important sign of depression for over a century(1,2). Although not contained in symptomatic criteria for major depressive disorder (MDD) in DSM-III(3), DSM-IIIR(4), DSM-IV(5), or DSM-5(6), they are found in 26 out of 28 detailed clinical descriptions of melancholia published from 1880-1900(1) and in 19 out of 21 of such descriptions of depression published in the 20th century(2). Given the current challenges in diagnosing MDD(2,7,8), where a large proportion of cases (ranging from 50% to 90%) remain untreated(9–11), the transformation of voice phenomena into diagnostic biomarkers could aid in both clinical and research arenas.

Clinical observations describe the speech patterns of depressed patients as slow, weak, low-pitched, and monotonous(1,2,12,13). These phenomena are typically quantified by increased pause time, lower volume, lower pitch, and reduced pitch variability(14–16). Many studies have sought to develop features from pitch as a biomarker for depression(17–26), but none have, to date, achieved sufficient accuracy and precision for clinical utility. The large number of both vocal features and confounds(24,25) imposes a multi-testing burden that requires large sample sizes which few studies have obtained(16). Also, a critical distinction between current mood and susceptibility to MDD on effects on voice has never been addressed(27–29). Furthermore, MDD is likely heterogeneous(30,31): studies not accounting for this may be underpowered (16).

Our study of the relationship between voice features and MDD was designed to be well-powered, using thousands of subjects, to be more robust to heterogeneity by analyzing cases with recurrent depression of one sex only, and able to separate susceptibility to MDD from the effect of current mood on voice features by using genetic data. We used a large case-control study of MDD where we could replicate findings in an independent sample, both from China. Since, of the four groups of speech features (source, spectral, prosodic, and formant features(16)), the association between depression and a key component of prosody, pitch, has repeatedly been observed(15,16), our analysis was restricted to examining pitch-related features. Our results return vocal features to a more central position in clinical and research work on MDD.

## Results

### Subjects

We used recordings conducted as part of the CONVERGE(32) (China, Oxford, and VCU Experimental Research on Genetic Epidemiology) study (3,968 cases and 4,354 controls). CONVERGE recruited only women with recurrent MDD from hospital settings and compared them with matched controls with no history of MDD, thus reducing heterogeneity in both genetic and vocal signals(33,34). A summary of demographic data from cases and controls is provided in **Table S1**; the relation of these to depression is reported in earlier publications (31,35–40). All recordings, obtained during diagnostic interviews, were listened to, and segments that contained only the patient’s voice at an adequate quality for the analyses (see the Method section in **Supplementary** for details) were identified. In this study, a “segment” refers to the longest continuous portion of an audio recording that contains only the patient’s voice, uninterrupted by other speakers. Segments are not split at pauses within the patient’s speech but are instead defined by changes in the speaker, ensuring that each segment represents uninterrupted speech from the patient alone. It can be as short as a single word or as long as a complex sentence or multiple sentences. This resulted in 364,929 voice segments with a duration greater than two seconds from 7,654 subjects. The selection of subjects for each component of the study is shown in **Figure S1**, which provides an overview of the design of the project (**Figure S1a**), and a PRISMA diagram to indicate how many cases and controls were discarded at different stages and pathways of analyses (**Figure S1b)**.

### Feature identification

The perceptual attribute of pitch corresponds to the physical measurement of fundamental frequency, or F0(15). Our interest in prosodic features of speech, particularly pitch (hereafter referred to as the F0) and change in pitch (ΔF0), led us to choose the INTERSPEECH 2016 Computational Paralinguistics Evaluation (COMPARE16) feature set (41,42). The primary application for the feature set has been depression detection(24,43–47) and it captures typical temporal information and long-term information either statically, through the use of utterance level statistics/functionals, or dynamically, through frame-based delta (ΔF0) coefficients, reflecting differences between the adjacent vectors’ feature coefficients(48,49). Its feature extraction process is well documented(50), with standardized and well-referenced methodologies that facilitate reproducibility and validation by other researchers in many languages (including Chinese)(51–54).

The COMPARE16 feature set contains 83 F0/ΔF0-based features, many of which are highly correlated (**Figure S2**). Implementing a feature selection process to remove redundancy (described in **Supplemental Methods**), we extracted 30 voice features (**Table S2**, their distributions are in **Figure S3** and **Figure S4**), providing a comprehensive characterization of the speaker’s prosodic patterns, pitch, and intonation(41,42) We calculated statistics and functions based on the time series of F0, and its differential values, namely pitch change speed (ΔF0). These statistics and functions include mean values, quartiles, range, and regression coefficients, which capture pitch trends and dynamics in speech.

### A two-stage meta-analysis identifies 20 features associated with MDD

Our association analysis had to account for a number of potential confounds. Not everyone in the study spoke the same language: 60% of the subjects spoke in standard Mandarin, whereas the rest spoke either local languages or Mandarin with local accents. This might not matter if language differences were randomized with respect to case status, but uneven case/control ratios between hospitals could confound the analysis. Similarly, the quality of the recordings varied, potentially confounding the association testing. While we made every attempt to ensure that the location of interviews was comparable (in outpatient departments) and that the interviews were carried out in the same way by clinically experienced interviewers that we had trained (Supplementary Methods), these and other, unknown confounders might impact the voice features.

We dealt with these issues as follows. First, during the process of identifying the patients’ voice segments, we annotated background noise and language. The noise was categorized into five levels, and a binary indicator tagged whether the subjects’ speech was in standard Mandarin or not. We included these features as covariates in our analyses.

Second, to account for differences between hospitals, we implemented a two-stage meta-analysis, in which associations were first calculated at the hospital level, including demographic features as covariates (**Table S1**), noise levels, and speech indicators, and subsequently pooling results using a random-effects model. We selected 27 hospitals with at least 100 individuals, yielding a total subject count of N = 5,681 (**Figure S1**). By analyzing cases and controls within each hospital first and then combining the results in a meta-analysis, we alleviated the risk of site-specific confounders. We identified 20 features significantly associated with MDD at a 5% FDR threshold (**Table 1**; a description of the vocal features is given in **Table S2**). All features were standardized (to give a standard deviation of 1) before analysis so that beta coefficients can be compared and interpreted. Eighteen features were significant under a family-wise error rate control using Bonferroni (P-value < 0.0017), with 14 features showing absolute β coefficients greater than 0.3. The most significantly associated feature is a ΔF0 measure (interquartile range; β = −1.07, SE = 0.07, P_FDR_ = 1.1 × 10^−49^).

**Table 1.** The table shows the names of the prosodic phenotypes, explained in Table S2. Beta values (Beta) P-values (P) and FDR corrected (FDR) are from the logistic regression analysis. Beta coefficients are derived from analyses of normalized voice features (with standard deviation of 1).Results from the CONVERGE study and an independently collected replication are shown. The column headed ‘Power’ shows the power of the replication study to detect the effect found in the discovery sample. The last three columns (‘Combined’) are results from a meta-analysis of CONVERGE and the Replication sample

| Features | Statistical Functionals | CONVERGE |  |  | Replication |  |  |  | Combined |  |  |
| --- | --- | --- | --- | --- | --- | --- | --- | --- | --- | --- | --- |
|  |  | Beta | P | P_FDR | Beta | P | P_FDR | Power | Beta | P | P_FDR |
| ΔF0_iqr1-3 | Interquartile range (3rd -1st) | -1.07 | 3.5E-51 | 1.1E-49 | -1.09 | 1.46E-07 | 0 | 1 | -1.07 | 3.4E-59 | 6.7E-58 |
| ΔF0_percentile99.0 | Maximum (99th percentile) | -0.97 | 1.2E-43 | 1.8E-42 | -0.97 | 5.35E-06 | 0 | 1 | -0.97 | 1.8E-49 | 1.8E-48 |
| F0_upleveltime90 | Time with F0>90th percentile | -0.80 | 1.5E-41 | 1.5E-40 | -0.78 | 5.37E-05 | 0.0001 | 1 | -0.80 | 1.9E-45 | 1.3E-44 |
| ΔF0_kurtosis | Kurtosis | 0.87 | 1.5E-39 | 1.1E-38 | 0.83 | 2.69E-04 | 0.0005 | 1 | 0.86 | 6.4E-42 | 3.2E-41 |
| ΔF0_rqmean | Root quadratic mean | -0.69 | 6.3E-31 | 3.8E-30 | -0.76 | 2.99E-04 | 0.0005 | 0.99 | -0.70 | 6.3E-34 | 2.1E-33 |
| ΔF0_quartile3 | 3rd quartile | -0.78 | 5.4E-30 | 2.7E-29 | -0.88 | 1.16E-05 | 0 | 0.98 | -0.80 | 4.1E-35 | 1.6E-34 |
| ΔF0_minPos | Position of the minimum | -0.60 | 3.3E-25 | 1.4E-24 | -0.57 | 7.05E-07 | 0 | 0.96 | -0.59 | 7.2E-31 | 2.0E-30 |
| ΔF0_amean | Mean | 0.43 | 5.8E-20 | 2.0E-19 | 0.56 | 5.85E-08 | 0 | 0.88 | 0.45 | 3.0E-26 | 7.4E-26 |
| ΔF0_linregc1 | Slope of linear regression | -0.39 | 6.0E-20 | 2.0E-19 | -0.55 | 3.12E-03 | 0.0042 | 0.88 | -0.41 | 8.7E-21 | 1.5E-20 |
| F0_range | Range | 0.57 | 1.0E-19 | 3.1E-19 | 0.48 | 2.35E-03 | 0.0034 | 0.88 | 0.55 | 3.7E-22 | 8.2E-22 |
| ΔF0_maxPos | Position of the maximum | -0.55 | 1.8E-19 | 4.9E-19 | -0.61 | 1.36E-03 | 0.0023 | 0.87 | -0.56 | 5.4E-22 | 1.1E-21 |
| ΔF0_qregc1 | 1st quadratic regression coefficient | 0.38 | 1.0E-17 | 2.6E-17 | 0.47 | 1.04E-04 | 0.0002 | 0.83 | 0.40 | 9.2E-22 | 1.7E-21 |
| ΔF0_flatness | Flatness | -0.43 | 5.5E-14 | 1.3E-13 | -0.39 | 2.85E-05 | 0.0001 | 0.68 | -0.42 | 6.6E-18 | 1.0E-17 |
| F0_lpc2 | 2nd linear prediction coding coefficient | 0.33 | 1.3E-09 | 2.7E-09 | 0.41 | 2.22E-05 | 0.0001 | 0.44 | 0.34 | 6.0E-13 | 8.6E-13 |
| F0_lpc4 | 4th linear prediction coding coefficient | -0.28 | 4.9E-09 | 9.8E-09 | -0.25 | 1.07E-01 | 0.1259 | 0.4 | -0.27 | 2.7E-09 | 3.6E-09 |
| F0_kurtosis | Kurtosis | 0.23 | 1.1E-05 | 2.0E-05 | 0.30 | 1.81E-03 | 0.0028 | 0.19 | 0.24 | 3.6E-08 | 4.5E-08 |
| F0_lpc0 | 0th linear prediction coding coefficient | -0.29 | 9.0E-05 | 1.6E-04 | -0.39 | 1.97E-02 | 0.0246 | 0.14 | -0.30 | 6.1E-06 | 7.2E-06 |
| ΔF0_risetime | Time with which ΔF0 is rising | -0.15 | 4.1E-04 | 6.9E-04 | -0.25 | 1.39E-01 | 0.1539 | 0.1 | -0.16 | 8.9E-05 | 9.9E-05 |
| F0_ff0_maxSegLen | Maximum length of voiced segments with F0 > 0 | 0.19 | 6.3E-03 | 1.0E-02 | 0.12 | 2.69E-01 | 0.2837 | 0.05 | 0.19 | 3.2E-03 | 3.4E-03 |
| F0_rqmean | Root quadratic mean | 0.14 | 9.0E-03 | 1.3E-02 | 0.00 | 9.97E-01 | 0.9974 | 0.05 | 0.12 | 1.7E-02 | 1.7E-02 |

To test the sensitivity of our results to differences between cases and controls, we compared analyses with and without the inclusion of 20 genetic principal components (PCs). The results of this analysis are presented in **Table S3**, and a comparison of the betas with and without adjusting for genetic PCs is presented in **Figure S5**. The correlation between betas in the two analyses was r = 0.99, P = 8.7 × 10^−28^. These results demonstrate a high degree of consistency in the estimated association effects, irrespective of the adjustment for genetic PCs. There is no overall decrease in significance with the inclusion of the genetic covariates, implying that cases and controls are overall adequately matched by location, which implicitly also means matching by accent.

### Replication in an independent sample

We evaluated the 20 associated features in an independent sample. The replication sample was collected six years after the discovery CONVERGE data, using the same selection criteria and interview protocol (described in **Supplemental Methods**). The replication sample collected data from three new hospitals, and one that was part of the CONVERGE sample. It used none of the same interviewers and the participants did not overlap with the discovery sample. A description of the sample is given in **Table S4**. While the replication sample is smaller than the discovery sample (1,084, **Figure S1**), power to detect twelve of the features was greater than 80% (**Table 1**). Again, we listened to all recordings, annotated them for quality and accent, extracted prosodic features, analyzed the association within each hospital, and combined results by meta-analysis. As **Table 1** shows, 14 features exceeded a Bonferroni corrected threshold of 0.0025 (0.05/20) and 16 exceeded an FDR 5% threshold. We argue that this strong replication finding in an independent sample, excludes systematic bias in the way recordings were made, the way interviews were conducted, and differences in accent among subjects and differences between hospitals.

Finally, we jointly analyzed the discovery and replication samples by meta-analysis and show the results in **Table 1**. Eighteen features exceeded the Bonferroni corrected threshold and all exceeded the 5% FDR threshold. The three most significantly associated features were ΔF0 interquartile range (β = −1.07, SE = 0.07, P_FDR_ = 6.8 × 10^−58^), which measures the range between the 25th and 75th percentile of pitch change speed (ΔF0), ΔF0 maximum (β = −0.97, SE = 0.07, P_FDR_ = 1.8 × 10^−48^), which measures the highest value of pitch change speed, and time with F0>90th percentile (β = −0.80, SE = 0.06, P_FDR_ = 1.3 × 10^−44^), which measures the amount of time the pitch stays above the 90th percentile of its range. We found that there was no significant heterogeneity in the association effects between Mandarin speakers and non-Mandarin speakers (**Supplemental Results**).

### Differences between case and control interviews do not account for the associations

We considered next one additional potential confound: the possible impact of the questions asked at interview. The interview for cases is typically more than twice as long as for controls, as we ask about past occurrences of depression and associated stressful life events. Could the emotion associated with this questioning alter speech in such a way as bias our findings? We conducted a sensitivity analysis on responses to neutral questions to check if the effects remain consistent across contexts.

We selected two questions from the demographic section of the interview based on their high response rates (**Table S5**) and neutral nature. These question (D2.A: “What is your date of birth?” and D10: “How much do you weigh while wearing indoor clothing?”) were chosen because they are unlikely to trigger emotional differences between MDD cases and controls. We identified 533 subjects with voice response to question D2.A and 617 to question D10. The average segment durations were 3.37 seconds (SD=3.05), and 8.47 seconds (SD=2.69), respectively. For each question, we used the corresponding segments to extract the 16 pitch features that were associated with MDD in our main analysis. Using the two-stage meta-analysis method again, we re-estimated their associations. Due to the small sample sizes, power to detect effects was low so we used a one-sided binomial sign test to test consistency in the direction of association effects between the two analyses.

The estimated association effects in context-constrained analysis are reported in **Table S6**. We found that for question D10, three out of 16 pitch features maintained significant associations with MDD at FDR<0.05. Remarkably, 15 out of 16 features showed the same direction of association effects, a fraction significantly higher than chance (Binomial P= 0.00026). For D2.A, despite the average duration being only 3.37 seconds, three features achieved nominal significance for associations (uncorrected P<0.05), and 12 out of 16 pitch features showed consistent directions of association effects (Binomial P= 0.038). In total, 12 out of 16 voice features showed consistent directions of association effects across all four analyses. We conclude that the findings from the main analyses are not biased by the context of the interview.

As a summary for these analyses, **Figure S6** shows the effect sizes (beta coefficients) and the 95% confidence intervals for the association between 16 voice F0/ΔF0 features and MDD in the discovery (CONVERGE), replication and single segment analyses.

### Genetic Correlations Between Pitch Features and MDD

Cases for the CONVERGE study were identified as those who have a history of recurrent MDD, and though all were ascertained through hospitals, many were in remission. This raises the important question as to what the association with voice features represents: does it reflect their current low mood, compared to controls, or does it reflect their history of MDD? We addressed this question in the following way.

To see if any voice features correlated with current mood, we used a standard assessment of current mood for subjects, the depressive symptom checklist (SCL)(55). These data were only available for the replication sample. The distributions of SCL scores for cases and controls are presented in **Figure S7**. Of the 16 pitch features, 11 showed a significant association with current depressive symptoms, after FDR correction (**Table 2**).

**Table 2.** Voice pitch features associated with SCL scores. The associations between pitch features and SCL scores were estimated in the replication sample using a two-stage meta-analysis. Asterisks indicate heritable features, from Table 3.

| **Feature** | **Beta** | **SE** | **P** | **P_FDR** |
| --- | --- | --- | --- | --- |
| $\Delta F0\_maxPos$ | -0.222 | 0.04 | 3.4E-08 | 5.5E-07 |
| $\Delta F0\_percentile99.0$ * | -0.296 | 0.074 | 6.7E-05 | 5.3E-04 |
| $\Delta F0\_rqmean$ | -0.238 | 0.067 | 3.6E-04 | 0.001 |
| $\Delta F0\_quartile3$ | -0.286 | 0.081 | 3.9E-04 | 0.001 |
| $F0\_upleveltime90$ | -0.246 | 0.066 | 1.9E-04 | 0.001 |
| $\Delta F0\_iqr1-3$ * | -0.309 | 0.097 | 0.001 | 0.004 |
| $\Delta F0\_kurtosis$ * | 0.225 | 0.081 | 0.005 | 0.01 |
| $F0\_lpc2$ | 0.118 | 0.042 | 0.005 | 0.01 |
| $\Delta F0\_amean$ | 0.133 | 0.053 | 0.01 | 0.02 |
| $F0\_kurtosis$ * | 0.097 | 0.041 | 0.02 | 0.03 |
| $F0\_range$ | 0.128 | 0.057 | 0.02 | 0.03 |
| $\Delta F0\_flatness$ | -0.116 | 0.056 | 0.04 | 0.05 |
| $\Delta F0\_qregc1$ | 0.104 | 0.062 | 0.09 | 0.12 |
| $\Delta F0\_linregc1$ | -0.103 | 0.067 | 0.12 | 0.14 |
| $\Delta F0\_minPos$ | -0.13 | 0.086 | 0.13 | 0.14 |
| $F0\_lpc0$ | -0.047 | 0.104 | 0.65 | 0.65 |

These results confirm that most of the features we found to be associated with MDD are correlated with current mood (the relatively smaller sample for this analysis cannot exclude the possibility that all features are thus associated). To examine whether the association reflected a genetic effect common to both variability in vocal features and susceptibility to MDD we estimated the SNP-based heritability for each of the 16 pitch features (this analysis was carried out with CONVERGE data, the only group for which there are genetic data(32)). Results are presented in **Table 3**. Four features were heritable at FDR<0.05. We repeated the heritability analyses adjusting for more genetic PCs to determine whether population structure might contribute to the correlation and found that the heritability remained significant even after adjusting for as many as 60 genetic PCs (**Table S7**). While we cannot rule out the possibility that all vocal features are to some extent heritable (our sample size is too small to confidently detect heritabilities of less than 10%), **Table 3** shows that SNP-based heritability varies significantly: the estimated confidence interval of the heritability for two, ΔF0_maxPos and ΔF0_qregc1, lie outside those for ΔF0_iqr1-3. The low heritability of these features indicate the genetic effects are unlikely to be the only contributing factor to the association with MDD. We did not find any genome-wide significant SNPs for these heritable features (**Supplemental Results**), presumably owing to the limited sample size.

**Table 3.** Heritable voice pitch features and their genetic correlation with MDD.

| Feature | SNP heritability |  |  |  | Genetic Correlation with MDD |  |  |  |
| --- | --- | --- | --- | --- | --- | --- | --- | --- |
|  | h2 | 95% CI | P | P_FDR | rg | 95% CI | P | P_FDR |
| **$\Delta F0\_iqr1-3$** | **0.171** | **(0.071, 0.272)** | **0.0004** | **0.006** | **-0.45** | **(-0.77, -0.13)** | **0.03** | **0.04** |
| **F0_kurtosis** | **0.134** | **(0.035, 0.234)** | **0.004** | **0.03** | -0.3 | (-1.14, 0.54) | 0.2 | 0.2 |
| **$\Delta F0\_kurtosis$** | **0.125** | **(0.025, 0.225)** | **0.007** | **0.03** | **0.55** | **(0.23, 0.88)** | **0.01** | **0.02** |
| **$\Delta F0\_percentile99.0$** | **0.121** | **(0.022, 0.221)** | **0.008** | **0.03** | **-0.7** | **(-1.28, -0.11)** | **4.2E-05** | **0.0002** |
| $\Delta F0\_flatness$ | 0.105 | (0.007, 0.203) | 0.02 | 0.05 | | | | |
| F0_lpc2 | 0.093 | (-0.005, 0.191) | 0.03 | 0.08 |  |  |  |  |
| $\Delta F0\_rqmean$ | 0.058 | (-0.040, 0.157) | 0.12 | 0.25 | | | | |
| F0_upleveltime90 | 0.044 | (-0.053, 0.141) | 0.18 | 0.32 |  |  |  |  |
| $\Delta F0\_minPos$ | 0.03 | (-0.067, 0.128) | 0.27 | 0.40 | | | | |
| $\Delta F0\_amean$ | 0.024 | (-0.073, 0.120) | 0.31 | 0.40 | | | | |
| $\Delta F0\_quartile3$ | 0.019 | (-0.078, 0.116) | 0.35 | 0.40 | | | | |
| F0_lpc0 | 0.019 | (-0.079, 0.116) | 0.35 | 0.40 |  |  |  |  |
| F0_range | 0.001 | (-0.096, 0.098) | 0.49 | 0.49 |  |  |  |  |
| $\Delta F0\_linregc2$ | -0.004 | (-0.101, 0.093) | 0.47 | 0.49 | | | | |
| $\Delta F0\_qregc1$ | -0.028 | (-0.124, 0.068) | 0.28 | 0.40 | | | | |
| $\Delta F0\_maxPos$ | -0.065 | (-0.160, 0.030) | 0.09 | 0.21 | | | | |

We estimated the genetic correlation with MDD for the four features with evidence of heritability and found that three ΔF0 features had significant genetic correlations (**Table 3**). They were: 1) the interquartile range (IQR1-3), quantifying the variation of speed in pitch change; 2) the kurtosis, signaling the extremity of speed in pitch change; and 3) the maximum, representing the speed of the fastest pitch change. There was no detectable genetic correlation with MDD for one heritable feature, ΔF0_kurtosis. Again, while we cannot exclude the possibility of some degree of genetic correlation for this and other features, our results indicate that genetic effects alone cannot explain the association with MDD for all features. The vocal features index a composite of heritable and non-heritable contributions to mood change.

### Associations Between Pitch Features and MDD Symptoms, Risk Factors, and Comorbidities

The deep set of phenotypes available in CONVERGE, which includes MDD symptoms, environmental risk factors, comorbid disease, and suicidality, permit us to explore other associations for the vocal features associated with MDD. For these exploratory analyses we included all 30 voice features and tested association with 30 traits (detailed in **Table S8**). The results of within-case two-stage meta-analysis are shown in **Table S8**. We categorized the traits into six classes (MDD symptoms, MDD clinical features, suicidal features, co-morbid psychiatric disease, neuroticism and stressful life events) as we were interested in determining the effects on these categories 98 associations are significant at an uncorrected 5% significance threshold (where 45 are expected by chance). Surprisingly, features assessing stressful life events showed the greatest enrichment of low P-values. After applying a Bonferroni correction for the 900 tests (P<0.05/900=5.5 × 10^−5^) five associations were significant, four for stressful life events and one for the personality trait neuroticism. Two features replicated (corrected threshold P<0.05/5=0.01). Both associations were between the total number of stressful life events and ΔF0 features, including the IQR1-3 of ΔF0 (16 hospitals in CONVERGE, total N=2,064, β = −0.21, SE = 0.03, uncorrected P = 1.9 × 10^−11^; four hospitals in the replication, total N=295, β = −0.20, SE = 0.07, uncorrected P = 0.0078) and maximum of ΔF0 (16 hospitals in CONVERGE, total N=2,064, β = −0.19, SE = 0.03, uncorrected P = 6.5 × 10^−10^ ; four hospitals in the replication, total N=295, β = −0.20, SE = 0.08, uncorrected P = 0.0074).

### Classification Performance

If the voice features are to have any clinical utility, they must not just be associated with MDD, but they must predict it accurately. We took advantage of access to our two independently collected samples (discovery and replication), using the discovery group for training data (n=7,654) and the replication sample (n=1,189) to test the classification performance.

We compared the classification performance of a full model against a null model. The null model was a logistic regression (LR) model trained on the covariates. We then trained a full model using the covariates together with the identified voice features in discovery, based on the same LR method. **Table 4** illustrates these comparisons. Integrating voice data significantly enhanced the predictive accuracy of our models. Adding voice features to the LR model increased the AUC-ROC from 0.70 to 0.83 and the accuracy from 0.63 to 0.76.

**Table 4.** Classification performance using the identified voice pitch features. LR, Logistic Regression. XGBoost, Extreme Gradient Boosting SVM, Support Vector Machine. MLP, Multi-layer Perceptron

| Model | Method | Feature | AUC-ROC | AUC-PR | Sensitivity | Specificity | F1 Score | Accuracy |
| --- | --- | --- | --- | --- | --- | --- | --- | --- |
| NULL | LR | Demographic Covariates | 0.70 | 0.62 | 0.75 | 0.53 | 0.63 | 0.62 |
| FULL | LR | Demographic Covariates + Voice | 0.83 | 0.77 | 0.81 | 0.70 | 0.73 | 0.75 |
|  | SVM | Demographic Covariates + Voice | 0.86 | 0.80 | 0.88 | 0.67 | 0.75 | 0.76 |
|  | MLP | Demographic Covariates + Voice | 0.85 | 0.80 | 0.86 | 0.69 | 0.75 | 0.76 |
|  | XGBoost | Demographic Covariates + Voice | 0.90 | 0.88 | 0.85 | 0.81 | 0.80 | 0.82 |

We then tested whether the classification results were robust to different machine learning methods. We evaluated this on three established methods suitable for our dataset, support vector machine (SVM), extreme gradient boosting (XGBoost), and multi-layer perceptron (MLP). Results improved prediction, with XGBoost delivering an AUC of 0.90 (sensitivity of 0.85, and specificity of 0.81). **Figure 1** plots the ROC curves for a null model (using covariates only to classify depression) and the four models using voice features. The precision-recall curve of these models are in **Figure S8**.

**Figure 1.**
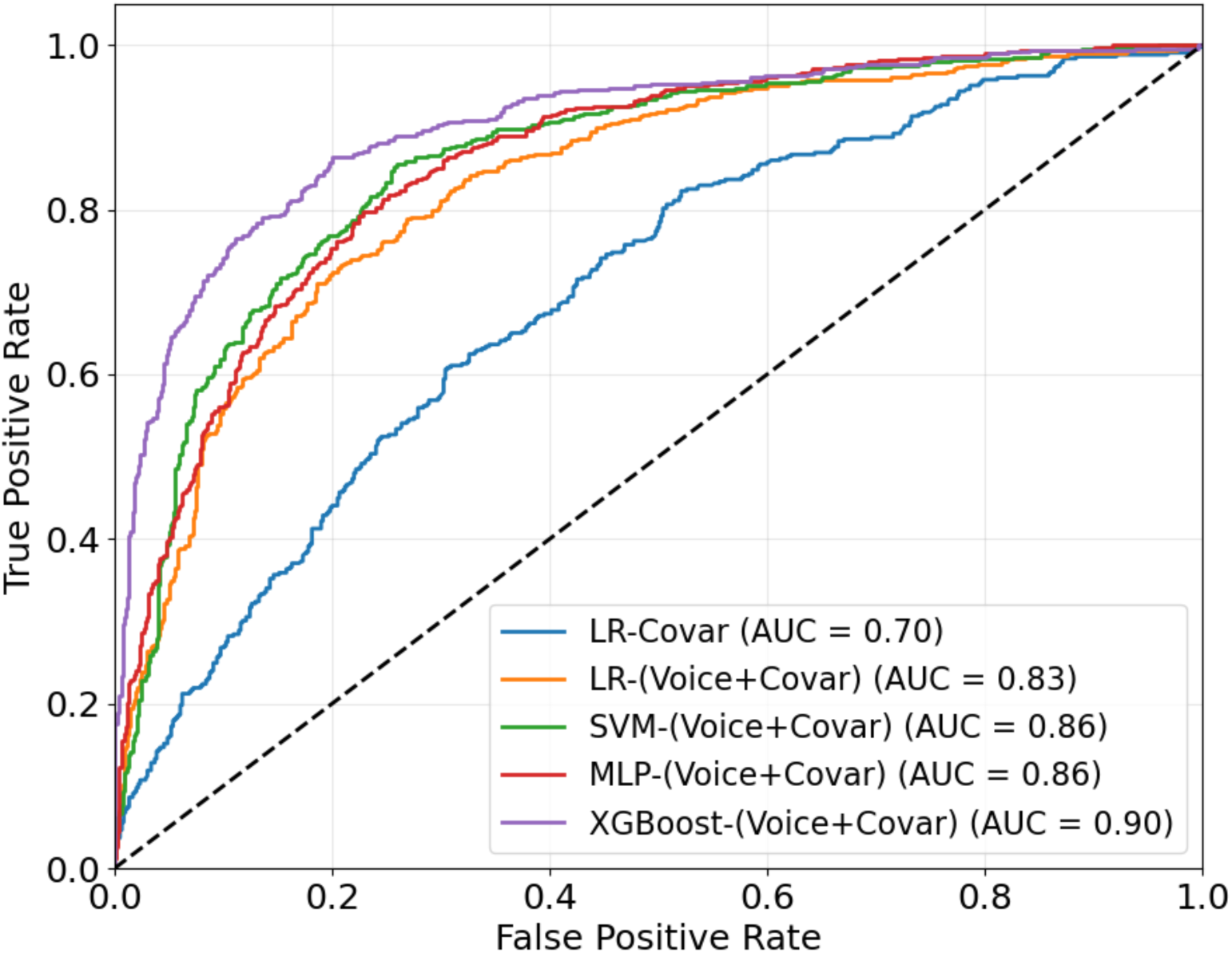
Receiver Operating Characteristic (ROC) Curve. The figure shows the ROC for four models for predicting depression from voice features, and a null model, a logistic regression model trained on demographic covariates only (LR-Covar). The full models are logistic regression (LR), support vector machine (SVM), multi-layer perceptron (MLP), and extreme gradient boosting (XGBoost), trained on voice and covariates (Covar+Voice). AUC: area under the curve.

## Discussion

We set out to find voice pitch features associated with MDD. By using a large and homogeneous case-control cohort, a two stage meta-analysis and an independent replication, we provide robust evidence that certain pitch features distinguished MDD cases from matched controls. The associated features were a slower change in pitch and a particularly uneven distribution of these variations. Features measuring the variability and extremity of pitch change speed were heritable and had genetic correlations with MDD, which we interpret to mean that at least some of the association between variation in pitch and susceptibility to depression is genetic in origin. Classification of those with and without depression, based on vocal features, was achieved with an AUC of 0.90, highlighting the potential use of these features as biomarkers for MDD detection and secondary prevention.

Establishing a robust, replicable association between voice features and MDD is difficult because of the numerous confounds that could potentially introduce systematic differences between cases and controls and thus corrupt our findings. We addressed this concern by using a large, and as far as possible homogeneous sample of depression. Our study used only women with recurrent MDD in a population where many comorbid disorders, such as smoking, alcohol, and drug abuse, are rare or practically non-existent(32). By adopting a two stage meta-analysis to take into account variation between hospitals, and using an independent replication sample, our results are unlikely to be explained by differences between hospitals, location, accent, quality of the recording or the interview questions.

Our findings support and extend previous studies which have indicated potential links between pitch patterns and MDD, but were limited by smaller sample sizes or more heterogeneous cohorts(15,16). First, our large sample size provided adequate power to test several pitch features from a standardized features set, providing more fine-grained quantitative evidence for the descriptions of the monotonous speech pattern in MDD than in previous studies. Previous studies have found that depressed people speak more slowly with lower pitch and decreased variability(16,25). Here, our study showed MDD was negatively associated with features measuring how fast pitch changes (the maximum, the 3rd quartile, and the root quadratic mean of ΔF0, **Table 1**), indicating that the reduced rate of change in pitch is a characteristic of voice in MDD patients. We also found that MDD patients spend less time in their upper vocal range (Time with F0>90^th^ percentile, **Table 1**), affirming the “low-pitched” pattern.

Second, our results indicate that MDD’s pitch dynamics involve more than reduced variability, showing a broader pitch range and more extreme values (range and kurtosis of F0, **Table 1**). Our analysis also revealed an uneven distribution of the speed with which an MDD patient’s pitch changes, as shown by the negative association between MDD and the flatness of ΔF0 and the positive association with the kurtosis of ΔF0 (**Table 1**). Overall, these various features enrich our understanding of pitch dynamics, demonstrating a pattern of slower change in pitch, yet with more frequent occurrences of extreme values and pitch change speed.

Third, our research examined the relationship between voice features and the effects of current low mood, and the effects of susceptibility to MDD. Some vocal features might be more reflective of a person’s underlying propensity towards developing MDD, while others could be more indicative of a current depressive state. We found that some vocal features were indeed heritable, but still correlated with changes in current mood: individuals with an increased genetic risk of MDD may have a smaller value of speed for the fastest pitch change, thus being unable to speak as fast as those without depression. They may show a narrower IQR of pitch change speeds and more frequently occurring extreme changes of pitch (higher kurtosis). Shared genetic effects exist between at least some ΔF0 features and MDD, but while our low power to detect heritability and genetic correlations raises the possibility that the other features may also be associated in this way, our findings are consistent with vocal features’ association with both current low mood and susceptibility to MDD.

We also found that two heritable voice features were associated with the number of stressful life events. The reason for these associations is unclear, but suggests the possibility that stressful life events reveal a latent predisposition to depression(56,57), evidenced through a change in vocal features.

It is interesting to consider physiological interpretations of our findings. Possibly, changes in pitch could reflect tiredness, or the psychomotor retardation that characterizes an episode of MDD. We do not have physiological assessments of our subjects that characterize these features (all of our data are from interviews, and therefore reflect the subjects’ perceptions) but it is worth pointing out that at some level physiological and psychological contributions will be confounded. For example retardation of thought (psychological) can result in a slowness of expression (motor effect), so that in many cases the distinction may not be relevant.

Could the features we identified provide clinically useful predictions? A key aim of our research was to find vocal biomarkers that could take on this role. Applying XGBoost to the independent test dataset we obtained an AUC-ROC of 0.90, and a level of accuracy indicating that the features could be useful in identifying cases of MDD. We applied three models for classification (SVM, XGBoost, and MLP) because relying on a single model would not provide sufficient evidence for the robustness and generalizability of the voice features across different machine learning approaches. SVM excels at handling high-dimensional data and constructing optimal decision boundaries, but it assumes that the data is linearly separable in the kernel-transformed feature space(58). If the relationship between the features and depression states is highly non-linear, SVM may struggle to find an optimal solution. In contrast, XGBoost(59), an ensemble of decision trees, can capture complex feature interactions and handle non-linear relationships. However, it may not be as effective as deep learning models like MLP in capturing hierarchical representations of the data. MLP, with its deep learning architecture, can learn intricate patterns and hierarchical representations, but it is prone to overfitting, particularly when the dataset is small or the network is overly complex(60). By applying the voice features and geographical covariates to all three models, we provide a robust justification for the usefulness and generalizability of the voice features in depression state classification.

Our results should be assessed with respect to several limitations. First, we only recruited Han Chinese women with recurrent MDD. Our results may not extrapolate to men, those with single episode MDD, or to non-Chinese speakers. Second, our analysis focused solely on pitch features, and future studies should explore other feature types. Neural network-based approaches, which can learn directly from raw audio signals, also hold promise for detecting depression but face challenges in portability and interpretability, particularly when addressing complex confounding factors(61). Third, although our context-constrained analysis demonstrates that the signals we found are persistent across speech content, we cannot separate pitch differences due to word choice from pitch differences due to emotional content without additional experiments directly controlling the linguistic context.

While we don’t know how far results will generalize outside the female Chinese cohort, the findings reveal that vocal features can be used to identify MDD cases with high accuracy and we expect that with improvements, such as the inclusion of additional voice features, even higher predictive accuracy may be obtainable. Our hope is that these findings will further encourage efforts to assess changes in the voice, long understood by experienced clinicians to be a valuable sign, returning it to a more central position in clinical and research work on MDD.

## Methods

### Participants

We used data from the CONVERGE(32) study, in which women with recurrent MDD were recruited from 58 provincial mental health centers and psychiatric departments of general medical hospitals in 45 cities and 23 provinces of China. Participants were aged between 30-60, with two or more episodes of MDD that met the DSM-IV criteria(5), with the first episode occurring between ages 14-50. Cases were excluded if they had pre-existing bipolar disorder, nonaffective psychosis, smoking/nicotine dependence (alcohol and substance abuse were virtually absent in this study, so it was not assessed), or mental retardation. Control subjects, screened to exclude a history of MDD, were recruited from patients undergoing minor surgical procedures at general hospitals and individuals attending local community centers. The replication study(43) used the same inclusion/exclusion criteria as CONVERGE, and recruited samples from 20 different hospitals in China, with a final sample size of 1,189 (**Figure S1**). This study was approved by Institutional Review Boards at UCLA, Bio-x Center, Shanghai Jiao Tong University (M16033), and local hospitals. All participants provided written informed consent.

### Data Collection

All subjects went through a semi-structured interview using a computerized assessment system as outlined previously(43) and described in **Supplementary Methods**. Recordings for cases were obtained in outpatient clinics. Controls were recorded in outpatient clinics and in community health centers. Recordings were not standardized and varied in quality and content. All participants provided DNA samples for genetic analysis. Details of DNA sequencing and genotype imputation have been previously reported(32) and described briefly in **Supplementary Methods**.

### Covariates

The covariates were five demographic variables and two recording quality variables. The demographic variables were age, education level, occupation, marital status, and social class. The recording quality referred to noise level and accent. The noise level and accent label were determined subjectively by the listeners during the process of identifying the patients’ voice segments. The noise was categorized into four levels: 1) No noise; 2) Slight noise but the subject’s speech was clear; 3) Noise present but the content of the subject’s speech could be clearly heard; 4) High noise levels and unclear speech. Note that noise level 4 means that the speech, although difficult to understand, can still be comprehended with extra effort. And samples were excluded during quality controls stage if the speech is not able to comprehended at all. The accent was a binary label that indicated whether the subjects’ speech was in standard Mandarin or not.

### Voice Data Preprocessing

8,322 subjects the subjects recruited in CONVERGE had interview recordings (**Figure S1**). To obtain the subjects’ utterances, a group of undergraduates listened to the recordings to identify any voice segments from the subjects with a duration >2 seconds. Audio samples were excluded where the noise was so prominent that the content of the subject’s speech could not be understood, which yielded 7,654 subjects with available segments. All segments from the same subject were concatenated in the order in which they occur in the interview and down sampled to 8 kHz. Two postgraduate psychological students listened to all the segments to ensure that no speech voice other than the subjects was included in the segments and that no words were cut off mid-way.

The preprocessing procedure in the replication study was the same as in CONVERGE. Of the initially recruited 1,301 participants (551 cases), 1,189 subjects (including 490 cases) had available voice segments (**Figure S1**). All segments from the same subject were concatenated into one, to extract voice features for replicating the association between voice and MDD and identifying the voice features associated with SCL (see distribution of the audio segment length in **Figure S9**). The speech segments in the replication study were further annotated to indicate the specific question that prompted each spoken response (**Table S5**). This additional level of data analysis was introduced to better understand the relationship between the interview content and the participants’ speech patterns. We additionally extracted the same voice features on the non-concatenated segments corresponding to a single question for a sensitivity analysis, which we referred to as, the single-item analysis (described below).

### Voice features

We used the INTERSPEECH 2016 Computational Paralinguistics Evaluation(41,42). Calculations were implemented in the openSMILE python package v2.4.2(62) and described in **Supplementary Methods**. Given that many of the features were highly correlated (for example, the arithmetic and root-quadratic mean of F0, as shown in **Figure S2**), we removed redundant features (described in **Supplementary Methods**), resulting in a set of 15 F0-based features and 15 ΔF0-based features. We provide in Supplemental **Table S2** technical definitions of the 30 features used, along with non-technical explanations of what each feature measures.

### Two-stage meta-analysis

We used a two stage meta-analytic framework to take into account differences between hospitals. In the first stage, for each hospital a linear regression model was fitted for each F0-related feature as the dependent variable using MDD and covariates as the predictor variables. We applied rank-based inverse normal transformation to the voice features. At stage 2, beta coefficients for MDD and standard errors from stage 1 were pooled using random-effects meta-analysis(63), assuming that the true effect sizes in different sites are not exactly the same but are drawn from a distribution of effect sizes. P-values were FDR-adjusted(64). In the second stage, we repeated the analyses in four hospitals with sample sizes ≥ 100 (N=1,084, **Figure S1**). We performed the same procedure as in the two-stage meta-analysis above. We combined results from both discovery and replication cohorts using random-effects meta-analysis(63).

### Heritability and Genetic Correlations

Heritability and genetic correlations were estimated on the 7,654 subjects in CONVERGE (**Figure S1**). The SNP-based heritability used a generalized REML (restricted maximum likelihood) method implemented in LDAK(65). We applied rank-based inverse normal transformation to the voice features and incorporated the above covariates and 20 genetic PCs. P-values were FDR-adjusted. For heritable voice features, we estimated their genetic correlation with MDD, adjusting for these same covariates and 20 genetic PCs. The genetic correlation was calculated through a bivariate GREML analysis implemented in GCTA(66,67). P-values were FDR-adjusted based on the number of heritable voice features.

### Associations between pitch features and current mood

To identify biomarkers for current mood, subjects in the replication cohort were given a 16-item, self-administered questionnaire assessing the severity of depression-related symptoms on a five-point distress scale over the past 30 days (subscales for depression symptom checklist, SCL)(55). We used the same two-stage meta-analysis method to estimate the association between the 16 voice features and SCL scores. All four hospitals from the replication cohort with sample sizes ≥ 100 were selected (N=1,084, **Figure S1**). At stage 1, for each hospital, a linear regression model was fitted for each pitch feature as the dependent variable using SCL scores and the covariates as the predictor variables. At stage 2, beta coefficients for SCL and standard errors from stage 1 were pooled using random-effects meta-analyses(63). SCL scores were standardized using rank-based inverse normal transformation. P-values were FDR-adjusted.

### Classification Model

We employed a logistic regression model using seven covariates (age, education level, occupation, marital status, social class, noise level, and accent) to establish a null model. Full models that incorporated both these covariates and voice features were then developed. We included all 20 voice features identified as associated with MDD during the discovery stage, including those not replicated. We used samples available in the discovery group for training data (n=7,654). For the test data set we used the replication sample (n=1,189). There was no re-estimation of weights in the test sample.

We compared the results of the full model with the null model based on logistic regression. Then we tested the classification performances using SVM, XGBoost, and MLP as the classifiers. The performance of models was evaluated across several metrics: accuracy, sensitivity, specificity, AUC-ROC, AUC-PR, and F1-score. To identify the best hyperparameters for each model, a grid search with 5-folds cross-validation was employed on the training dataset. The above process was implemented in python with package scikit-learn(68) v1.2.2 and xgboost(59) v2.0.3.

### Associations Between Pitch Features and MDD Symptoms, Risk Factors, and Comorbidities

We examined the relationship between the 30 voice F0/ΔF0 features with 33 variables related to MDD, including eight risk factor variables, 11 comorbidity variables, seven symptoms, three variables about suicidality, age of onset, number of MDD episodes, neuroticism, and premenstrual syndrome score (summarized in **Table S8**). We again employed the two-stage meta-analysis procedure. At stage 1, for each hospital, a multivariate linear regression model was fitted for each pitch feature as the dependent variable using one of the above variables and covariates as the independent variables. At stage 2, we used the Q statistics to measure the heterogeneity of the pooled beta coefficients and standard errors. If the heterogeneity test is significant (P<0.05), we then used the random-effects model for meta-analyses, otherwise we used fixed-effects(64). We applied a Bonferroni correction to obtain a 5% significance threshold. Due to the high endorsement rates for certain variables within some hospitals, the hospitals included in the meta-analysis varied depending on the variable being analyzed (for example, if all cases from one hospital did not have suicidal attempts, this hospital would be excluded for the analysis of suicidal attempts at stage 1.). We reported the number of hospitals and sample size for each association along with the meta-results.

### Context-Constrained Analysis

We counted the total number of available voice segments for each question in the replication cohort and selected the two most frequently answered questions from the demographic section of the interview: D2.A (“What is your date of birth?”) and D10 (“How much do you weigh while wearing indoor clothing?”). For each question, we used the corresponding segments to extract the 16 voice features that were associated with MDD in our previous analysis. Finally, we re-assessed the associations between these voice features and MDD through the two-stage meta-analysis method. The limited voice duration and small sample size reduced power to detect a significant signal. We applied the one-sided binomial sign test to determine whether the number of voice features demonstrating consistent directions of association effects between the concatenated segments and the context-constrained segments was greater than expected by chance (that is, a one-sided test of whether this fraction is greater than 0.5).

## Data Availability

All the code used for analyses, site-level summary statistics necessary for generating the final meta-analysis results, along with the labels and model predictions needed to reproduce the ROC and precision-recall curves are available online at Figshare[https://doi.org/10.6084/m9.figshare.27165387.v2]. GWAS Summary Statistics are available at https://doi.org/10.6084/m9.figshare.24571321.v1

https://doi.org/10.6084/m9.figshare.24571321.v1

## Acknowledgments

This work was funded by NIH grant MH-122596

