## Supplemental Text and Figures for "Unraveling the Associations Between Voice Pitch and Major Depressive Disorder: A Multisite Genetic Study"

The Supplementary contained the following contents:

- Supplementary Methods
- Figure S1-S11
- Tables S1-S10 (in a separate MS Excel file)

**Supplementary Results**

*Analysis on accent (Mandarin vs. non-Mandarin)*

We carried out within-group analyses (of Mandarin speakers and non-Mandarin speakers) and compared the results by using Cochran’s Q test (1) for heterogeneity. The results are shown in the following **Table S9** and **Figure S10**. We found that there was no significant heterogeneity in the association effects between Mandarin speakers and non-Mandarin speakers. The lowest heterogeneity p value is 0.047 (for ΔF0_quartile3). This is the only P value that is <0.05 for heterogeneity test on the 20 features (after applying a multiple-test correction this result is not significant at a 5% level).

*Genome-wide association analysis (GWAS) on the heritable voice features*

We performed GWAS on the 7,654 subjects in CONVERGE (**Figure S1**) for the four heritable features in **Table 3**: ΔF0_iqr1-3, F0_kurtosis, ΔF0_kurtosis, and ΔF0_percentile99.0. We did not find any significant hits, presumably owing to the limited sample size. The Manhattan plots and Q-Q plots are in **Figure S11.** SNPs associated with corresponding features at a P-value ${<10}^{-5}$ are listed in **Table S10**.

GWAS Summary Statistics are available at https://doi.org/10.6084/m9.figshare.24571321.v1).

**Supplementary Methods**

*Interview protocol*

All subjects were interviewed using a computerized assessment system. Interviewers were postgraduate medical students, junior psychiatrists, or senior nurses, trained by the CONVERGE team for a minimum of 1 week. Interviews were recorded and the research team listened to at least two interviews from each interviewer to identify any errors in the way questions were asked and answers were interpreted. The interview protocol acquired the following assessments for psychopathology: i) CIDI (WHO 1997) section of MDD expanded to include a “deep” assessment of the DSM-IV A criteria for MDD, symptoms of DSM-IV melancholia, Beck’s cognitive triad (helplessness, hopefulness, and worthlessness), and irritability/anxiety; ii) CIDI section on dysthymia; iii) sections from interviews in the Virginia Adult Twin Study of Psychiatric and Substance Use Disorders (VATSPSUD) (2) for generalized anxiety disorder, panic, and five phobia subtypes (agoraphobia and social, situational, animal, and blood injury phobias), (iv) brief assessments of premenstrual syndrome and postnatal depression (3, 4), and (v) assessments of smoking/nicotine dependence (alcohol and substance abuse were virtually absent in this study, so it was not assessed). Additionally, four key environmental exposures known to be strongly associated with the risk of MDD were assessed in cases and controls: i) child sexual abuse; ii) parent-child relationships; iii) social support; and iv) stressful live events. Neuroticism was assessed using the full 23-item Eysenck personality questionnaire N scale. Family history of MDD was individually assessed in parents and full siblings using the Family History Research Diagnostic Criteria. In each case, measures used are those developed, field-tested, and validated in the VATSPSUD studies (4).

The interview protocol in the replication study mirrored that used in the CONVERGE study, with the addition of a 16-item, self-administered questionnaire assessing the severity of depression-related symptoms on a five-point distress scale over the past 30 days (subscales for depression in symptom checklist, SCL).

### *DNA Sequencing and Genotype imputation*

The CONVERGE study used low-coverage sequencing to genotype the sample. DNA was extracted from saliva samples using the Oragene protocol. Sequencing reads obtained from Illumina Hiseq machines were aligned to Genome Reference Consortium Human Build 37 patch release 5 (GRCh37.p5) with Stampy (v1.0.17)(5) using default parameters after filtering out reads containing adaptor sequences or consisting of more than 50% poor quality bases. The aligned reads were indexed, and PCR duplicates were marked for removal. Base quality score recalibration (BQSR) was performed on the BAM files using the Genome Analysis Toolkit (GATK) (6), with known SNPs and INDELs masked. Whole-genome sequences were acquired to a mean depth of 1.73 (95% confidence intervals (CIs) 0.7–4.3) per individual, from which 32,781,340 SNP sites were identified.

Variant discovery and genotyping were conducted using the GATK's UnifiedGenotyper, targeting polymorphic SNPs in the 1000 Genomes Project Phase 1 East Asian reference panel. The dbSNP v137 rsids were used to fill in the variant ID column of the output variant call format (VCF) files. A sensitivity threshold of 90% to SNPs in the 1000G Phase1 ASN panel was applied for SNP selection for imputation. This gave a total of 21,356,798 (9,053,391 known in 1000 Genomes Phase 1 ASN Panel and 11,486,024 novel) biallelic SNPs identified from all chromosomes and unassembled contigs. Genotype likelihoods were calculated using SNPtools (7). The imputation process was conducted using BEAGLE software (8).

*Feature selection process to remove redundant features*

From the COMPARE16 feature set, we obtained 83 F0/ΔF0-based features. We excluded one feature, the minimal length for F0 >0 because the values of this feature were the same for almost all subjects. They were forced to the frame size unit, according to reference (9). Next, given that many of the features were highly correlated (for example, the arithmetic and root-quadratic mean of F0) (**Figure S2**), we implemented a feature selection process to remove redundant features. We first calculated the pairwise Pearson correlation for each voice feature pair and ranked the voice features from high to low according to their sum of squared correlation r values with all other features. We then retained the first (leading) feature and excluded all other features that had an absolute r value >0.5 with the leading one. This process was repeated, each time keeping the next leading feature remaining in the list and excluding its correlates. The above procedure resulted in a set of 30 F0/ΔF0-based features (**Table S2**) that were representative of the original 82 features and not highly correlated with each other.

*Genome-Wide Association Studies (GWAS)*

We performed GWAS for each one of the heritable voice features on the 7,654 subjects utilizing the LDAK tool (10). A genetic relationship matrix (GRM), constructed from the genotype dataset, was utilized to correct for relatedness among the samples. We applied rank-based inverse normal transformation to the voice features and incorporated these covariates into our analysis: 20 genetic PCs, age, education level, occupation, marital status, social class, noise level, and accent.

**Supplementary Figures**

**Figure S1. Study overview.** a) study aims and main analyses. QC: quality control. The criteria of inclusion of sites for meta-analysis is a sample size N > 100, and a case/controls ratio between 0.1 and 0.9. b) PRISMA diagram of the quality control process and selection of sites for meta-analysis.

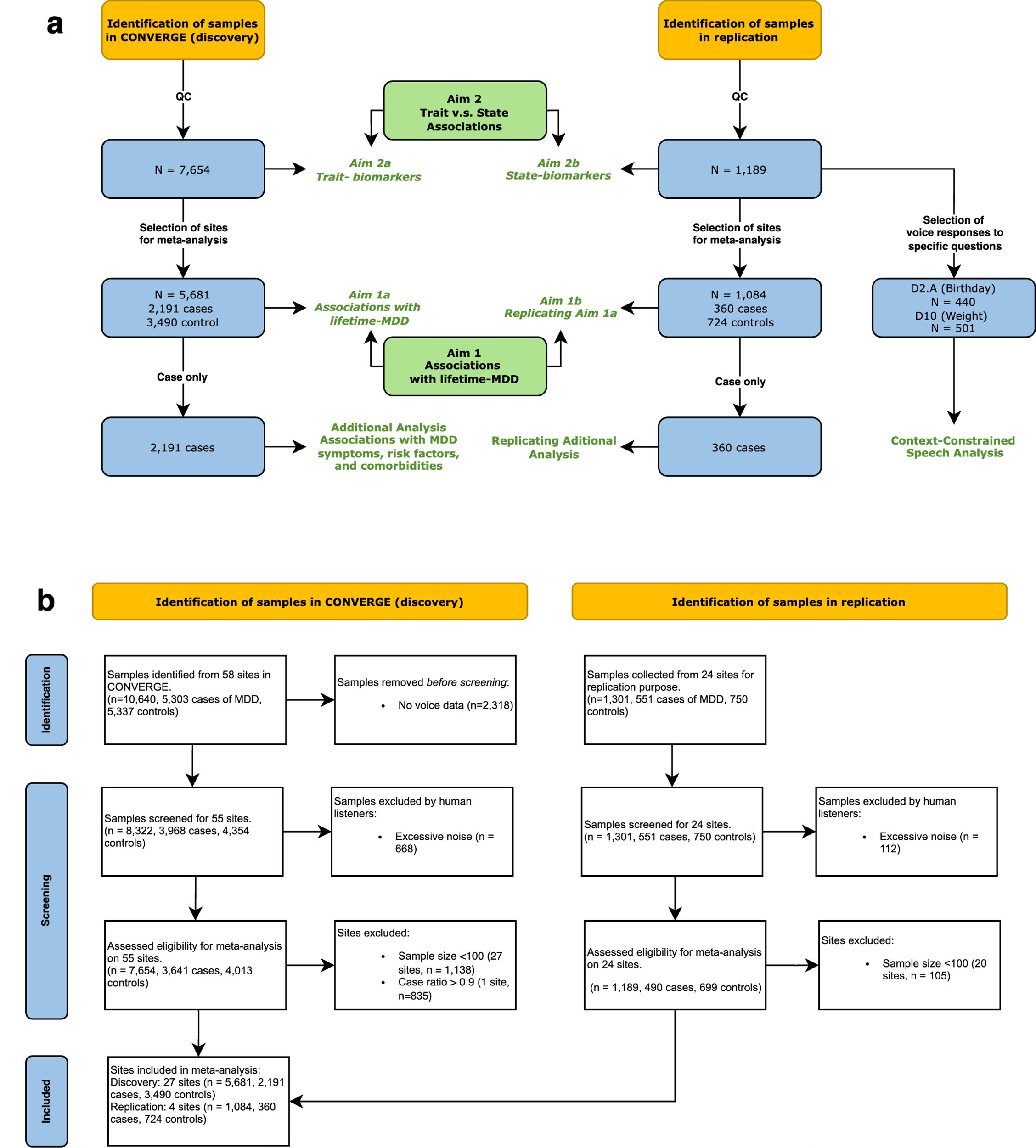

**Figure S2. Heatmap of the correlation r-values between the leading features and excluded features.**

The x-axis shows the 52 voice features excluded because of their high correlation with the 30 leading features (y-axis).

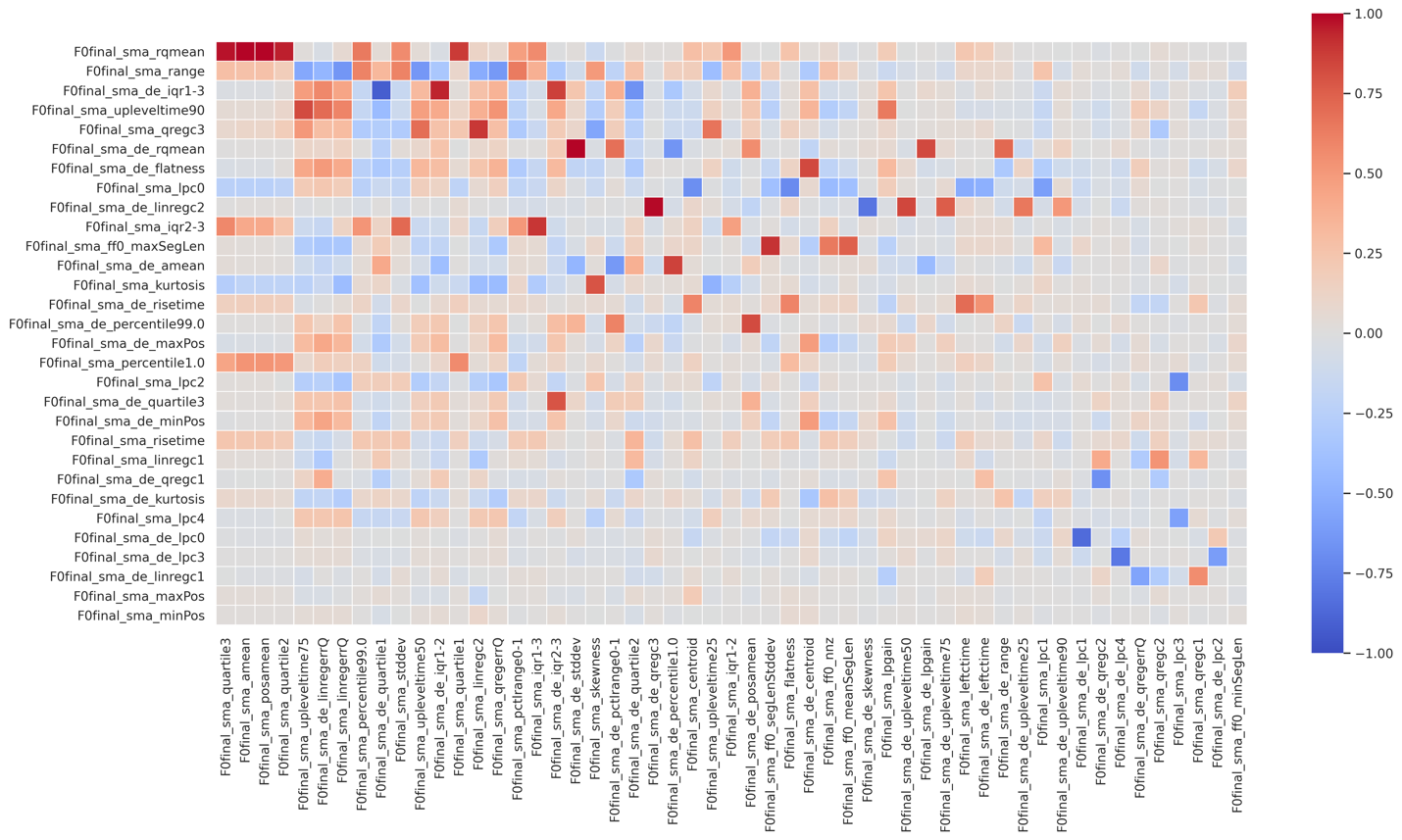

**Figure S3. Distribution of the 30 pitch features (original values).**

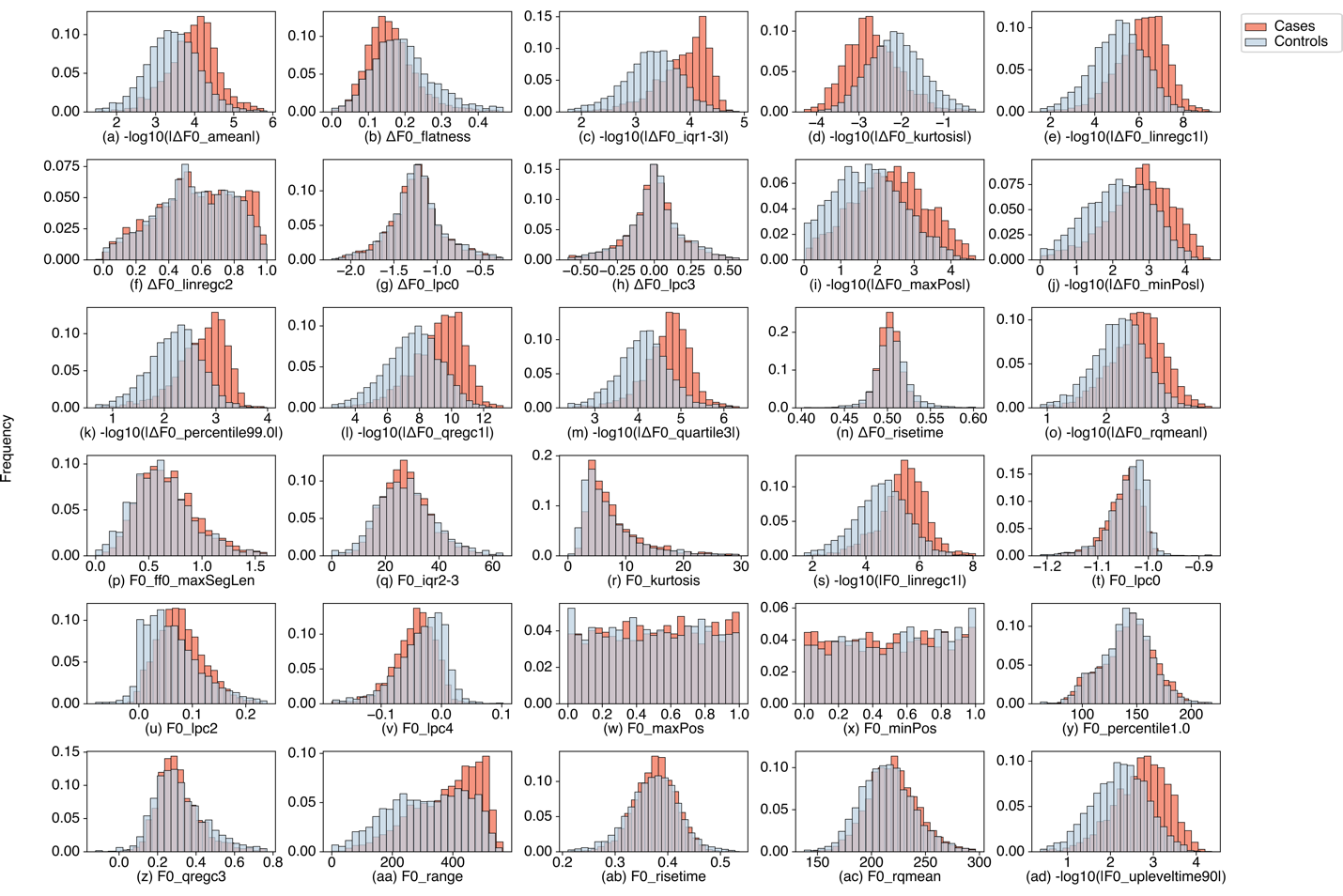

**Figure S4. Distribution of the 30 pitch features after normalization.**

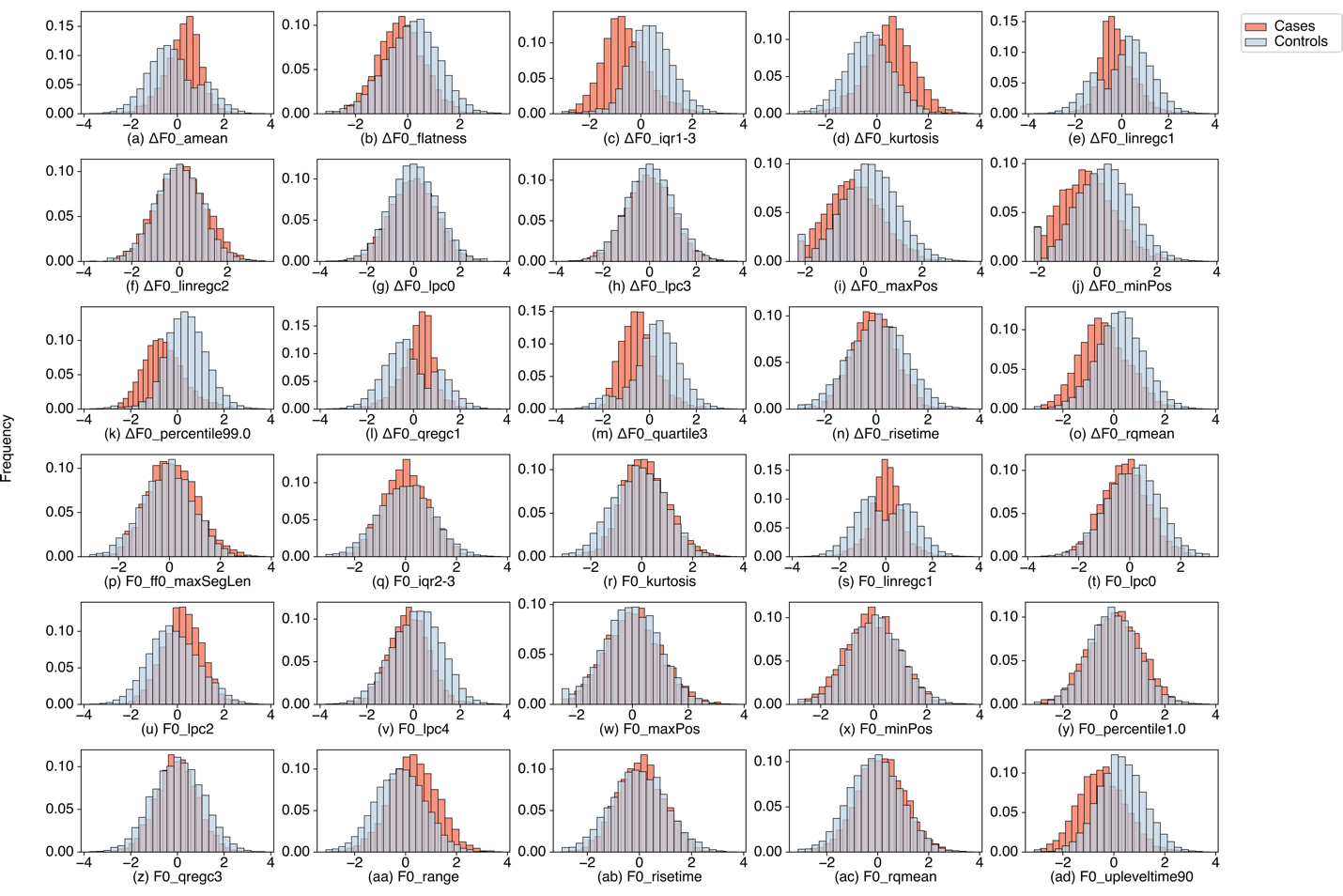

**Figure S5**. **Results of the meta-analysis with and without adjusting for genetic PCs.**

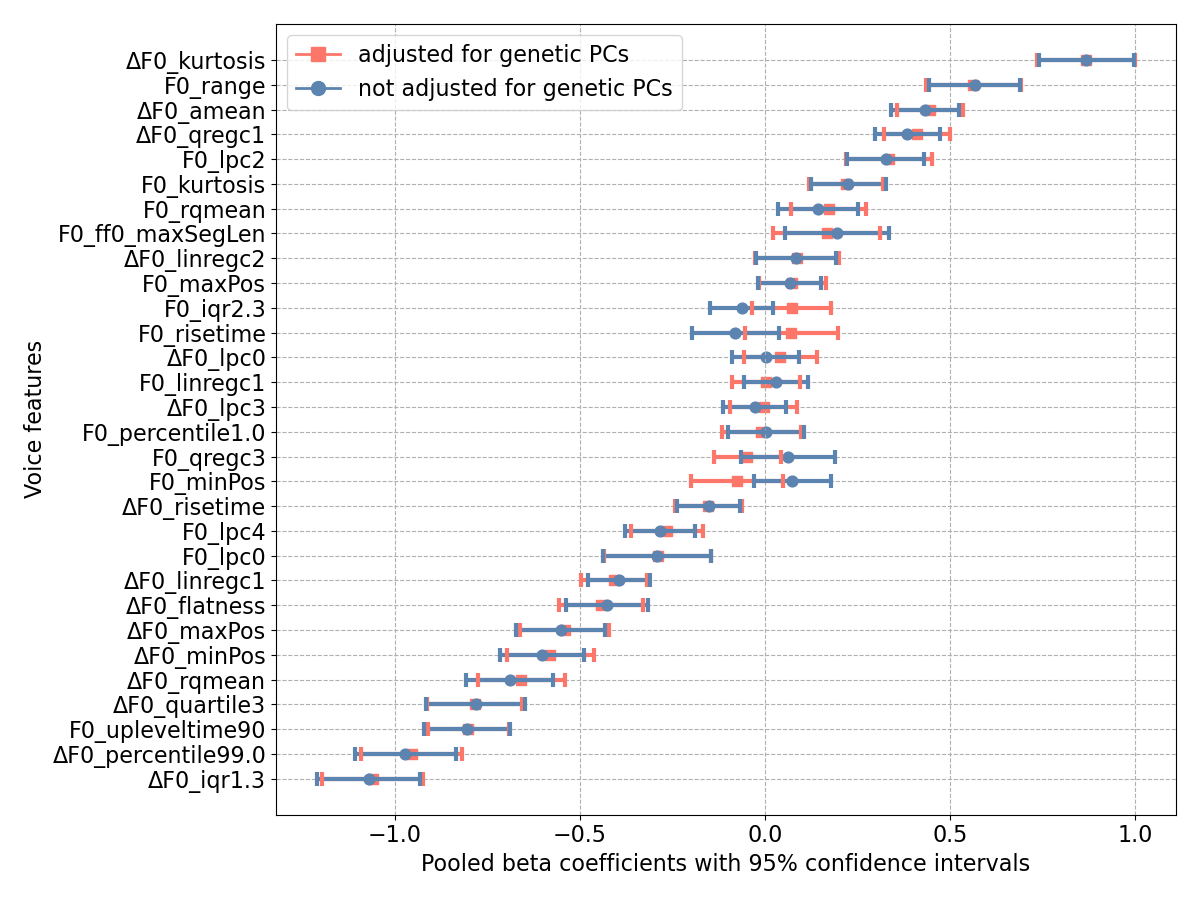

**Figure S6. Estimated beta coefficients with 95% confidence interval from associations between 16 voice F0/ΔF0 features and MDD.**

Estimations were conducted using the two-stage meta-analysis method on four cohorts: CONVERGE, replication, single-segment about weight (D10) from replication, and single-segment about birthday (D2.A) from replication.

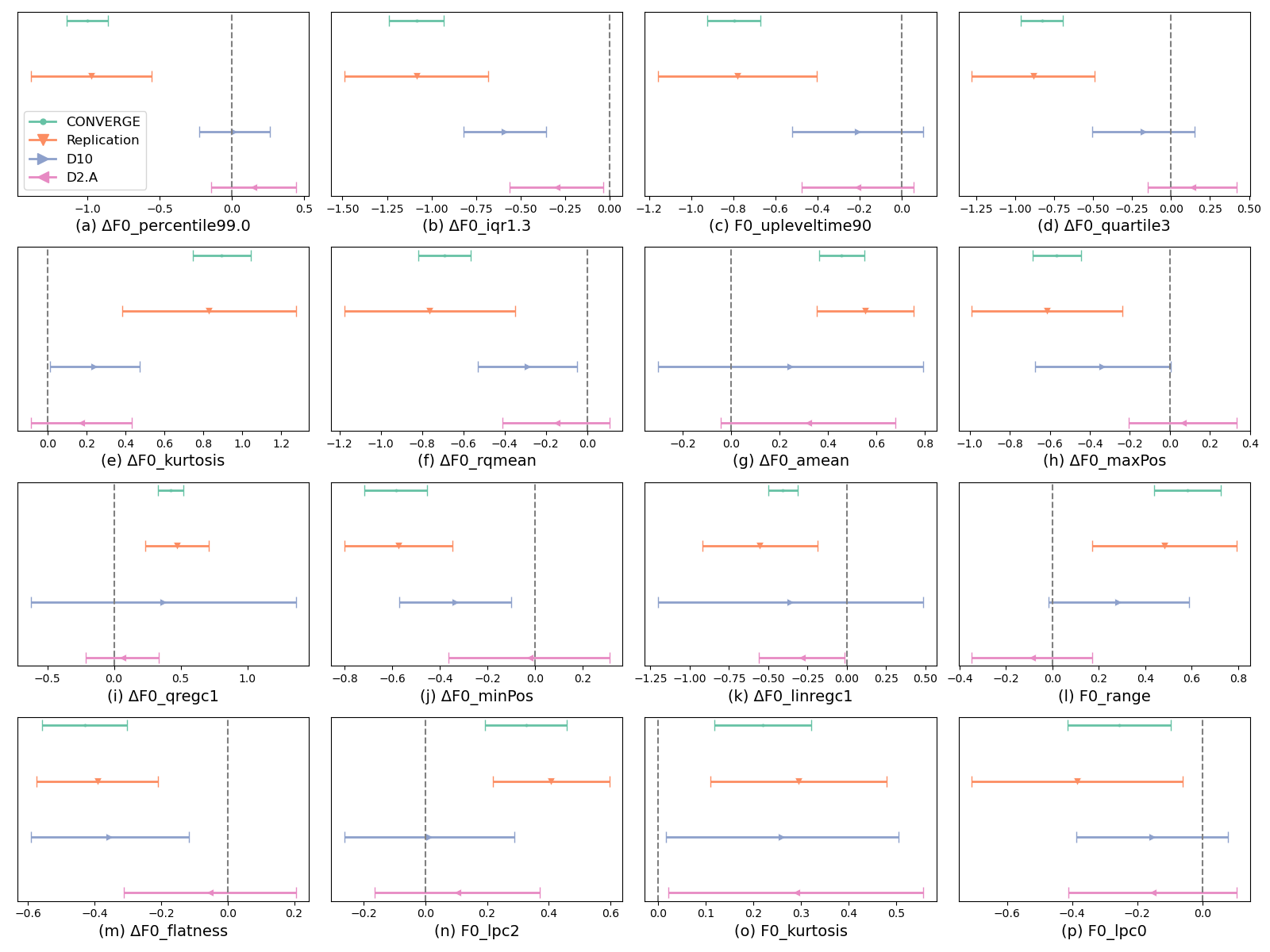

**Figure S7. The distribution of the Symptom Checklist (SCL) scores in MDD cases and controls.**

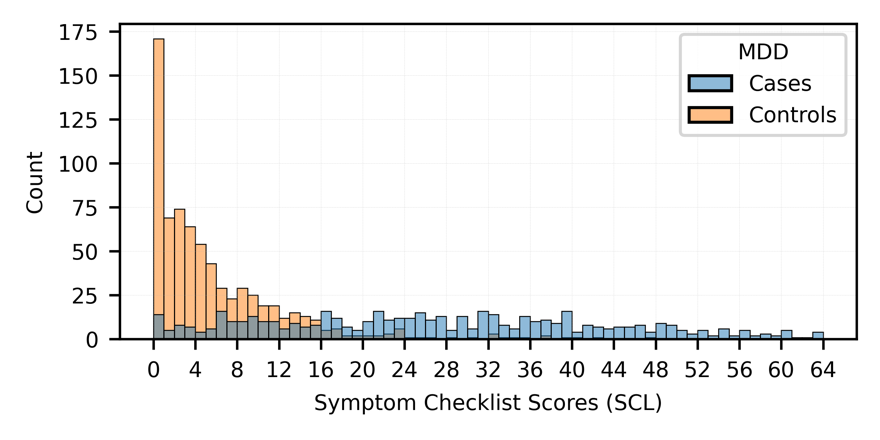

**Figure S8. Precision-recall curve of classification performance on test (replication) samples.** The figure shows the precison-recall curve for four models for predicting depression from voice features, and a null model, a logistic regression model trained on demographic covariates only (LR-Covar). The full models are logistic regression (LR), support vector machine (SVM), multi-layer perceptron (MLP), and extreme gradient boosting (XGBoost), trained on voice and covariates (Covar+Voice). AP: average precision score.

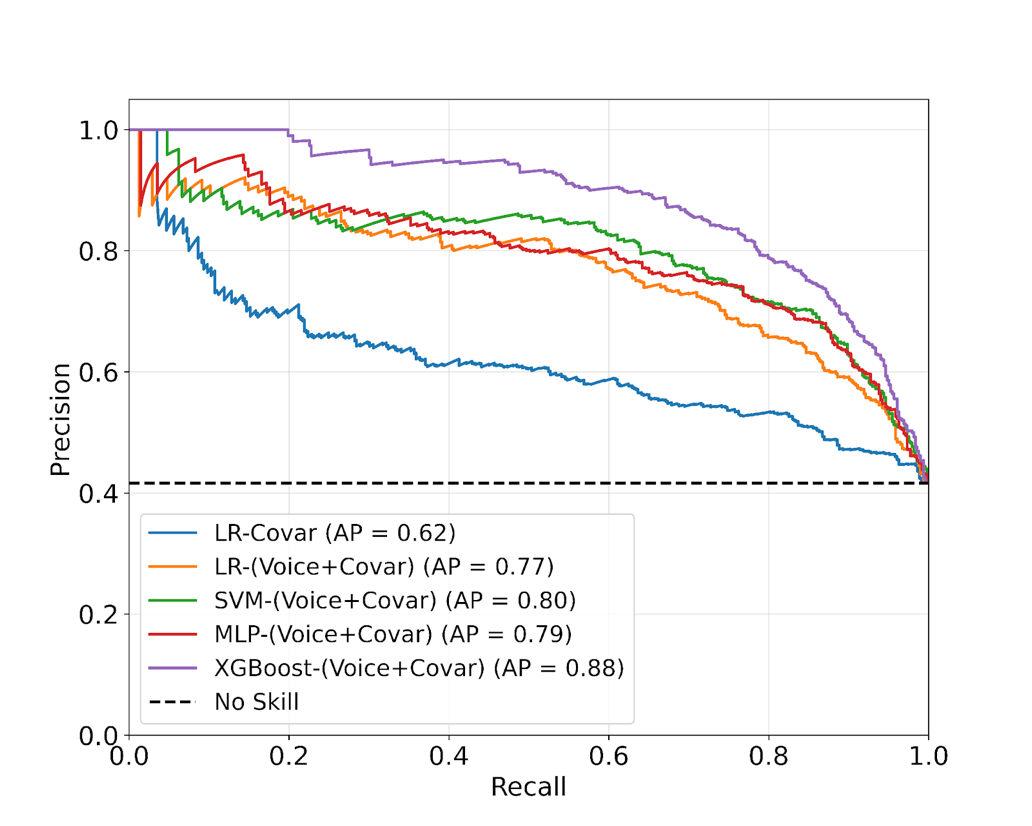

**Figure S9.** **Distribution of the audio segment length.**

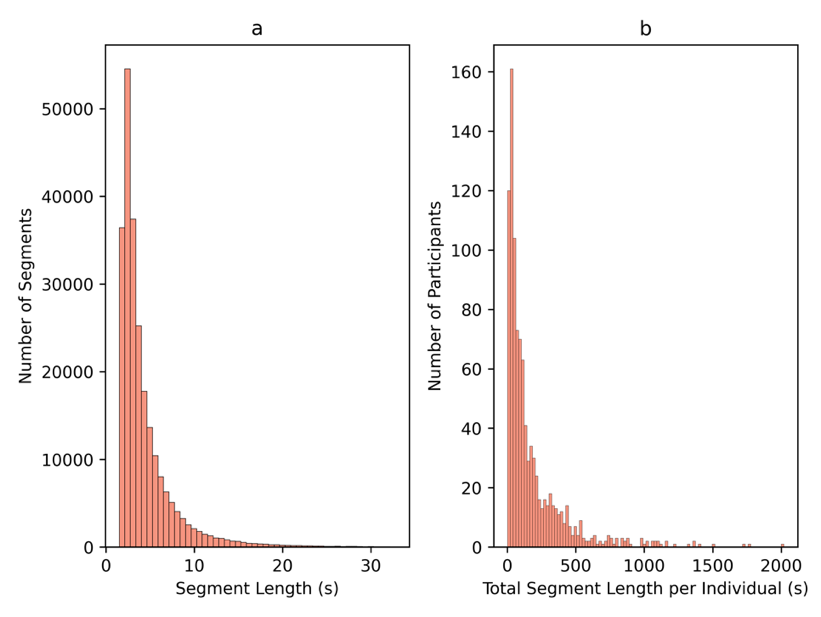

**Figure S10. Results of the meta-analysis on Mandarin and non-Mandarin speakers.**

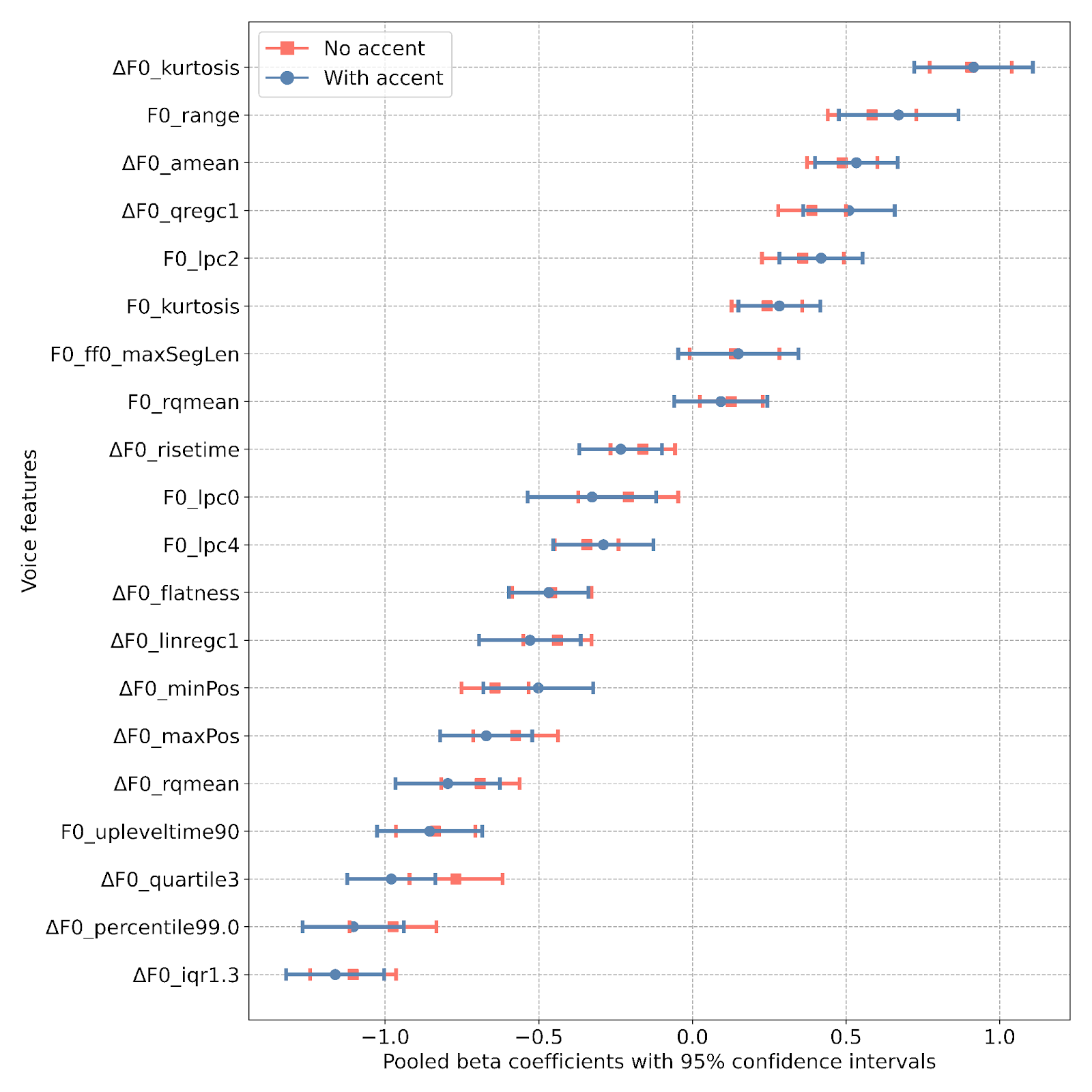

**Figure S11. GWAS results for a) ΔF0_iqr1-3, b) ΔF0_kurtosis, c) ΔF0_percentile99.0, and d) F0_kurtosis.**

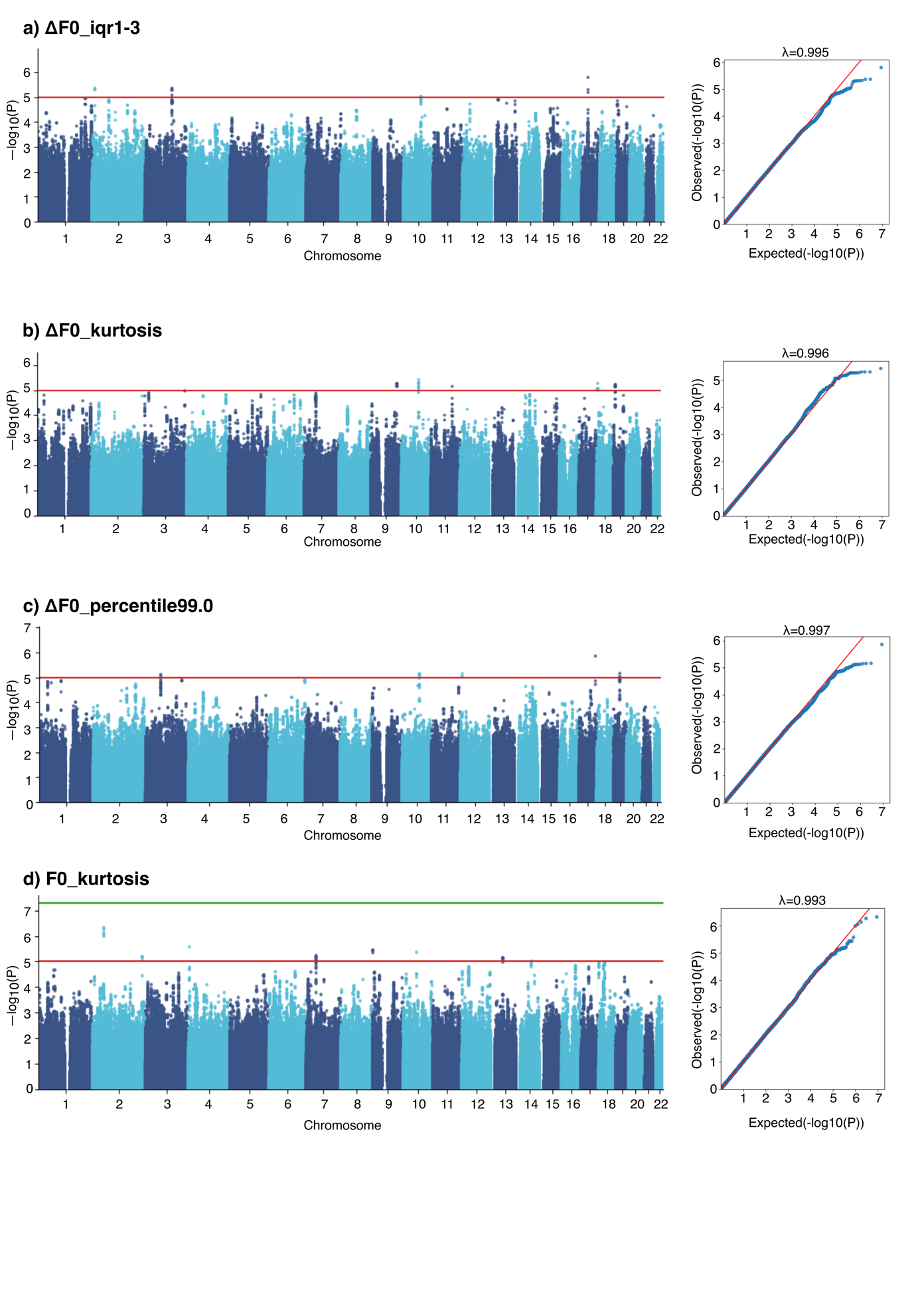
